# Stroke Care in Rural Kenya: The Gap Between Clinical Practice and Stroke Care Guidelines in Kiambu County, Kenya

**DOI:** 10.64898/2026.10.09.26365112

**Authors:** Elijah Yulu, Romeo Warera Ngesa, Samwel Maina Gatimu, Peter Kuria Waweru

**Author notes:** Correspondence: Romeo Ngesa.

## Abstract

**Background:** In Kenya, stroke is one of the top five causes of mortality, with a high rate of inpatient mortality. The American Heart Association (AHA) and American Stroke Association (ASA) translated scientific evidence into evidence-based clinical practice guidelines aimed at reducing inpatient mortality and improving the quality of care for stroke patients. Despite the proven benefits of the ASA stroke guidelines in improving stroke care, translation of the guidelines in various settings remains a key challenge. This study aimed to quantify the gap between evidence-based recommendations and current stroke care practices in a peri-urban referral facility in Kiambu, Kenya. The study was conducted at xxxxxxxxxx Hospital, a peri-urban referral facility in Kiambu County, Kenya.

**Methods:** We reviewed records of stroke patients attending xxxxxxxxxxx from January 1, 2020, to December 31, 2023. The study evaluated acute stroke management practices and secondary prevention against the 2019 American Heart Association/American Stroke Association (AHA/ASA) Guidelines for the Early Management of Acute Ischemic Stroke.

**Results:** The median distance travelled by stroke patients to the facility was 47.8 kilometres; further, the median onset-to-presentation time was 24 hours. The study evaluated eight clinical practices against the ASA guidelines. The facility lacked a dedicated stroke unit. Remarkably, patients arriving within the therapeutic window for intravenous alteplase and thrombectomy did not receive these treatments. Secondary prevention measures, including discharge on an antiplatelet regimen, were moderately adhered to; among the 86 eligible ischemic stroke patients, 67 (77.9%) were correctly discharged on aspirin. Similarly, 84/123 (68.3%) of the patients were discharged on statin therapy.

**Conclusion:** Evidence-based stroke care recommendations were minimally adhered to at this peri-urban referral facility in Kiambu, Kenya.

**Contribution:** This study quantifies the gap between evidence-based stroke care recommendations and current clinical practice in a peri-urban referral facility in Kenya, providing context-specific evidence to inform quality improvement initiatives and guideline implementation efforts in similar resource-constrained settings.

## Introduction

Stroke is the second leading cause of mortality globally with attributable deaths of more than six million (1,2). More than two-thirds of the stroke mortalities occur in developing countries, with sub-Saharan Africa (SSA) having the highest incidence, prevalence, and fatality from stroke (3). Compared to high-income regions, strokes in SSA occur in younger patients (4), resulting in significant disability rates in African settings, worsening the socioeconomic balance, and compounding the limitation of already scarce resources (5). In Kenya, stroke is among the top five leading causes of mortality (6).

A stroke is an abrupt onset of a focal or generalised neurological deficit of vascular origin lasting longer than 24 hours or leading to death (7). The main stroke subtypes are ischaemic stroke, intracerebral haemorrhage and subarachnoid haemorrhage (8). Ischaemic stroke is the commonest subtype, accounting for 87% of all strokes (9).

The commonest risk factors are hypertension, diabetes, hyperlipidaemia, high body mass index, and chronic kidney disease, accounting for up to two-thirds of the global stroke burden (10). Other factors thought to predispose to the growing burden of stroke include malnutrition, air pollution, changes in population dynamics, and the established role of communicable diseases (4).

Delayed presentation, mostly due to unawareness of stroke symptoms, is associated with poor outcomes and higher mortality (11). The signs and symptoms of stroke include sudden confusion, trouble with speech, trouble with sight, and trouble walking (12). A clinical study in Northwest Ethiopia identified hemiparesis (57.7%), loss of consciousness (20.7%), and aphasia (9%) as the most common presentations among stroke patients (13). Access to quality stroke care for stroke patients, including neurosurgical services for haemorrhagic stroke patients in SSA, is very restricted, especially in rural areas (14,15). Furthermore, the region is plagued by a lack of stroke specialists, necessary infrastructure, and essential thrombolytics for ischemic strokes, and endovascular/surgical interventions. A large proportion of stroke patients end up lying helpless in general beds, with bed sores, in hospitals with limited inpatient rehabilitation and no long-term rehabilitation (16). In Kenya, the in-hospital mortality for stroke patients is about one in four (25%) in the first month and about a third by the third month (17,18).

The American Heart Association and American Stroke Association (ASA) translated scientific evidence-based clinical practice guidelines aimed at reducing inpatient mortality, limiting neurological damage, and improving long-term functional independence for stroke patients. Based on high-quality evidence, these guidelines provide recommendations in rapid emergency triage, timely neuroimaging, acute reperfusion therapies including thrombolysis and thrombectomy, and the early initiation of secondary prevention medications, including antiplatelets, statins, and anticoagulants for stroke patients (19). Despite the proven benefits of the ASA stroke guidelines in improving stroke care, translation of the guidelines in various settings remains a key challenge. This study aims to quantify the gap between evidence-based recommendations and the current stroke care practices in a peri-urban referral facility in Kiambu, Kenya.

## Methods

We conducted a retrospective study to analyse the medical records at xxxxxxxx Hospital. We reviewed records of stroke patients attending the facility from January 1, 2020, to December 31, 2023. xxxxxxxxxxxxx Hospital is a 363-bed tertiary referral hospital located in Kiambu County, Kenya (20).The hospital is equipped with neuroimaging, vascular, and cardiac imaging facilities necessary for accurate diagnosis and phenotyping of strokes.

The study evaluated acute stroke management practices and secondary prevention against the 2019 American Heart Association/American Stroke Association Guidelines for the Early Management of Acute Ischemic Stroke. The study population included all patients (>18 years) with a diagnosis of a first-ever stroke who attended care at the study centre between January 2020 and December 2023. Eligible patients were adults over 18 years, with neuroimaging-confirmed stroke, which was defined as the abrupt onset of focal or generalised neurological deficit of vascular origin lasting for 24 hours or leading to death. Patient records with no scan and under the age of 18 years were excluded from the study. All consecutive patient records of individuals meeting the eligibility criteria who attended Hospital within 10 days of symptom onset during 2020–2023 were included in the retrospective analysis. Data extraction was carried out by two stroke specialists, an endovascular neurosurgeon and a family physician, from 19th October 2023 to 20^th^ March 2025.

Data were extracted retrospectively using a structured data collection tool. The tool was modified from the World Health Organisation’s STEPwise approach to surveillance (STEPS) instrument (21). Data were collected on sociodemographic information, pre-existing comorbidities, initial vital signs and laboratory tests, and time from presentation (door) to investigations, including neuroimaging. Treatment and outcome data, including admission to the intensive care unit, length of hospital stay, discharge medications (antihypertensives, antiplatelets, statins), and discharge status (alive or deceased), were also collected. Patient Identity information was accessible during data extraction; however, it was subsequently anonymised during analysis.

Due to the limitations of retrospective data, the audit only assessed eight key elements of the AHA/ASA guidelines. Furthermore, the Glasgow Coma Scale (GCS) was used as a proxy for the National Institutes of Health Stroke Scale (NIHSS) in place of a standardized severity score. Compliance for the eight evaluated components was operationalized using strict denominator criteria. The component evaluated included compliance with imaging, IV thrombolysis administration within 4.5 hours, thrombectomy for eligible patients, stroke unit availability, monitoring hyperglycaemia, and secondary prevention measures: the prescription of antiplatelets for non-cardioembolic strokes, and high-intensity statins for patients aged ≤75 years.

Data were cleaned and analysed using STATA version 17. The analysis included descriptive statistics where frequencies, means, modes, and medians were generated to summarize continuous variables. Median and interquartile range, frequency, cumulative frequency, frequency distribution tables, and cluster bar graphs were used to describe the sample population. Ethical approval for the study was obtained from the xxxxxxxxxx Hospital’s Ethics Review Committee (KH/ISERC/02718/0076/2023). The patient’s data was anonymised using numbers only.

**Table 1.** Operationalisation of variables.

| Variable | Description |
| --- | --- |
| Sociodemographic factors | Age: >45 years, <45 years |
|  | Gender: male, female |
|  | Residence: distance to facility in kilometres (km) |
| Medical history | Self-reported history of systemic hypertension |
|  | Self-reported history of diabetes mellitus |
| Access to health care services | Onset to presentation: time (in hours) from the onset of symptoms to presentation to the facility |
| Initial assessments (at baseline) | Blood pressure (mmHg) |
|  | Random blood sugar (mmol/l) |
|  | Glycated haemoglobin (HbA1C, %) |
|  | Haemoglobin level (mg/dl) |
| Severity of neurological impairment | Baseline and discharge assessment of neurological disability using Glasgow Coma Score (GCS): severe, moderate, mild |
|  | Neuroimaging (including lesion localisation, confirmation of stroke subtypes) |
| Stroke aetiology | Defined by stroke subtypes (ischemic, intracerebral haemorrhage, subarachnoid haemorrhage) |
| Discharge medications | Discharge on antihypertensives (yes/no) |
|  | Discharge on statins (yes/no) |
|  | Discharge on clopidogrel (yes/no) |
| Quality of life after stroke | Ambulation status on review (yes/no) |

## Results

### Patient characteristics

Among the 123 patient records analysed, 52 (42.3%) were male, and 71 were female (57.7%). The mean age of the patients was 66 years. The median distance travelled to the facility was 47.8 kilometres; further, the median onset to presentation time was 24 hours. About one-third of the patients were diabetic, and 60.2% (n=74) were known hypertensives. Remarkably, 19.5% (n=24) were unaware of their hypertensive status.

### Stroke subtypes

The commonest stroke subtype was ischemic stroke (n=86, 72.9%), followed by intracerebral haemorrhage (n=28, 23.7%), subarachnoid haemorrhage (n=3, 2.5%)—one patient presented with cerebral venous sinus thrombosis (n=1, 0.9%).

Three-quarters (74.4%) of ischemic stroke cases were partial anterior circulation infarcts, mainly involving the middle cerebral artery territory. Lacunar infarcts comprised 12% and posterior circulation strokes (11%), with the least being total anterior circulation strokes. The majority of intracerebral haemorrhages were deep bleeds (hypertensive; 62.5%), and 37.5% were lobar.

### Clinical presentation

On clinical assessment, 86.2% of the patients had hemiparesis, while only a quarter (n=31) of the patients had loss of consciousness. Lastly, some 45.5% (n=56) presented with a deviated mouth.

Table 1

**Table 1.** Operationalisation of variables.

| **Variable** | **Overall (N = 123)** |
| --- | --- |
| Age, years |  |
| Mean (SD, Range) | 66.1 (17 – 103) |
| **Sex, n (%)** |  |
| Female | 71 (57.7%) |
| Male | 52 (42.3%) |
| **Distance to facility, km** |  |
| Median (IQR) | 47.8 (26.7 – 71.6) |
| **Onset to presentation, hours** | N=115/123 |
| Median (IQR) | 24 (24 - 96) |
| **Baseline Comorbidities, n (%)** |  |
| **Diabetes** |  |
| yes | 39 (31.7%) |
| no | 84 (68.3%) |
| **Hypertension (HTN)** |  |
| Known Hypertensive | 74 (60.2%) |
| Unaware/ Newly diagnosed | 24 (19.5%) |
| Not hypertensive | 25 (20.3%) |
| **Clinical Presentation at Admission** |  |
| **stroke type** |  |
| Ischemic | 86 (69.9%) |
| ICH | 28 (22.8%) |
| Subarachnoid | 3 (2.4%) |
| CVST | 1 (0.8%) |
| Missing | 5 (4.1%) |
| **ischemic subtypes** |  |
| LACI | 10 (12.2%) |
| PACI | 61 (74.4%) |
| POCI | 9 (11.0%) |
| TACI | 2 (2.4%) |
| **ICH subtypes** |  |
| ICH-Deep | 15 (62.5%) |
| ICH-Lobar | 9 (37.5%) |
| **Neurological Deficits, n (%)** |  |
| **hemiparesis** |  |
| yes | 106 (86.2%) |
| no | 16 (13.0%) |
| missing | 1 (0.8%) |
| **mouth deviation** |  |
| yes | 56 (45.5%) |
| no | 66 (53.7%) |
| missing | 1 (0.8%) |
| **loss of consciousness** |  |
| yes | 31 (25.2%) |
| no | 90 (73.2%) |
| Missing | 2 (1.6%) |

The study evaluated eight clinical practices against the ASA guidelines [Table 2]. The ASA guidelines recommend that stroke patients be treated in a dedicated stroke unit with a multidisciplinary team; however, since the facility lacks a stroke unit, stroke patients are managed in the general wards. Despite the NIHSS being the preferred standardized severity measure, it was not used in the hospital. The GCS score was the documented stroke severity scale for 91.9% of the patients. Across the entire stroke cohort, brain imaging was performed in 97.6% of suspected stroke patients.

Despite five ischemic stroke patients arriving within the 4.5-hour window for IV thrombolysis administration, further evaluation revealed that none of the patients had IV alteplase records in the treatment sheet. Similarly, none of the patients qualifying for thrombectomy underwent mechanical thrombectomy or referred for that service.

Secondary prevention measures, including discharge on an antiplatelet regimen, were moderately adhered to; among the 86 eligible ischemic stroke patients, 67 (77.9%) were correctly discharged on aspirin. Similarly, 84/123 (68.3%) of the patients were discharged on statin therapy.

### Adherence to AHA/ASA Stroke performance measures

| **Specific ASA Guideline / Recommendation** | **Guideline Indicator** | **Compliance**<br>**(Yes/No/NA)** |
| --- | --- | --- |
|  |  | **N=123**<br>**Compliant, (%)** |
| Stroke Team: An acute stroke team (MDs, nurses, lab, radiology) is designated and available. | data not recorded |  |
| Door-to-Needle Time: Systems are in place to initiate IV alteplase within 60 minutes of hospital arrival. | data not recorded |  |
| Imaging Timing: Brain imaging is performed within 20 minutes of arrival. | data not recorded |  |
| Stroke Scale: A standardized severity scale (preferably NIHSS) is performed and documented. | Stroke severity documented (GCS recorded) | **113/123 (91.9%)** |
| Initial Imaging: All suspected stroke patients receive emergency brain imaging (NCCT or MRI) immediately upon arrival. | Brain imaging was performed for suspected stroke | **120/123 (97.6%)** |
| Vessel Imaging: Noninvasive intracranial vessel imaging (CTA/MRA) is recommended during initial evaluation for thrombectomy candidates. | data not recorded |  |
| Labs: Only blood glucose assessment must precede IV alteplase; do not delay for other labs (unless bleeding diathesis suspected). | No thrombolysis |  |
| IV alteplase is administered to eligible patients within 4.5 hours of symptom onset. | IV alteplase records in the treatment sheet | 0/5 (0%) |
| Thrombectomy is performed for eligible patients | Thrombectomy records in the patient's file. 20% of patients presenting within 24hrs | 0/63 (0%) |
| Stroke Unit: Patients are treated in a dedicated stroke unit with a multidisciplinary team. | No stroke Unit | 0/123 (0%) |
| Temperature: Fever (>38°C) is identified and treated with antipyretics. | data not recorded |  |
| Hyperglycemia: Blood glucose is monitored and treated to achieve a range of 140–180 mg/dL. | Blood glucose measured at admission | **107/123 (87.0%)** |
| Dysphagia Screen: A swallowing screen is performed before any oral intake (meds, food, liquid) to prevent pneumonia. | data not recorded |  |
| Nutrition: Enteral diet (tube feeding if needed) is started within 7 days of admission. | data not recorded |  |
| DVT Prophylaxis: Intermittent pneumatic compression (IPC) is used for immobile patients (elastic stockings are not recommended). | data not recorded |  |
| Depression: A structured depression inventory is administered to screen for post-stroke depression. | data not recorded |  |
| Rehabilitation: Early rehabilitation (but not high-intensity within 24h) is provided in an organized stroke care environment. | Data not recorded |  |
| Early Antiplatelets: Aspirin (160-325 mg) is given within 24-48 hours (if no IV alteplase given). | Discharged on antiplatelet therapy for ischemic type | **Aspirin**<br>**67/86 (77.9%)** |
|  | Discharged on antiplatelet therapy of ischemic type | **Clopidogrel**<br>**12/86 (14.0%)** |
| Atrial Fibrillation: Oral anticoagulation is initiated between 4–14 days after onset for AFib patients. | data not recorded |  |
| Statins: High-intensity statin therapy is initiated for patients $\leq 75$ years with clinical ASCVD (target LDL reduction $\geq 50\%$). | Discharged on statin therapy | 84/123 (68.3%) |
| Smoking: Smokers receive in-hospital counseling and nicotine replacement therapy (NRT). | data not recorded |  |
| Education: Patient/caregivers receive education on stroke risk factors, warning signs, and medications before discharge. | data not recorded |  |

### Stroke treatment and outcomes

A sensitivity analysis was done between the non-admitted (n=48) and admitted (n=67) groups, which revealed a statistically significant difference in clinical severity upon presentation (p=0.002). Patients not admitted had milder stroke as per the GCS scores, with 86.7% presenting with a Glasgow Coma Scale (GCS) score of 13–15, compared to 56.3% in the admitted cohort. These findings reveal a major challenge in bed capacity, whereby beds are preserved for only severe stroke patients. However, the comparison did not reveal statistical significance in sex, age, stroke type, or time to presentation between those admitted and those not admitted.

**Table 3.**
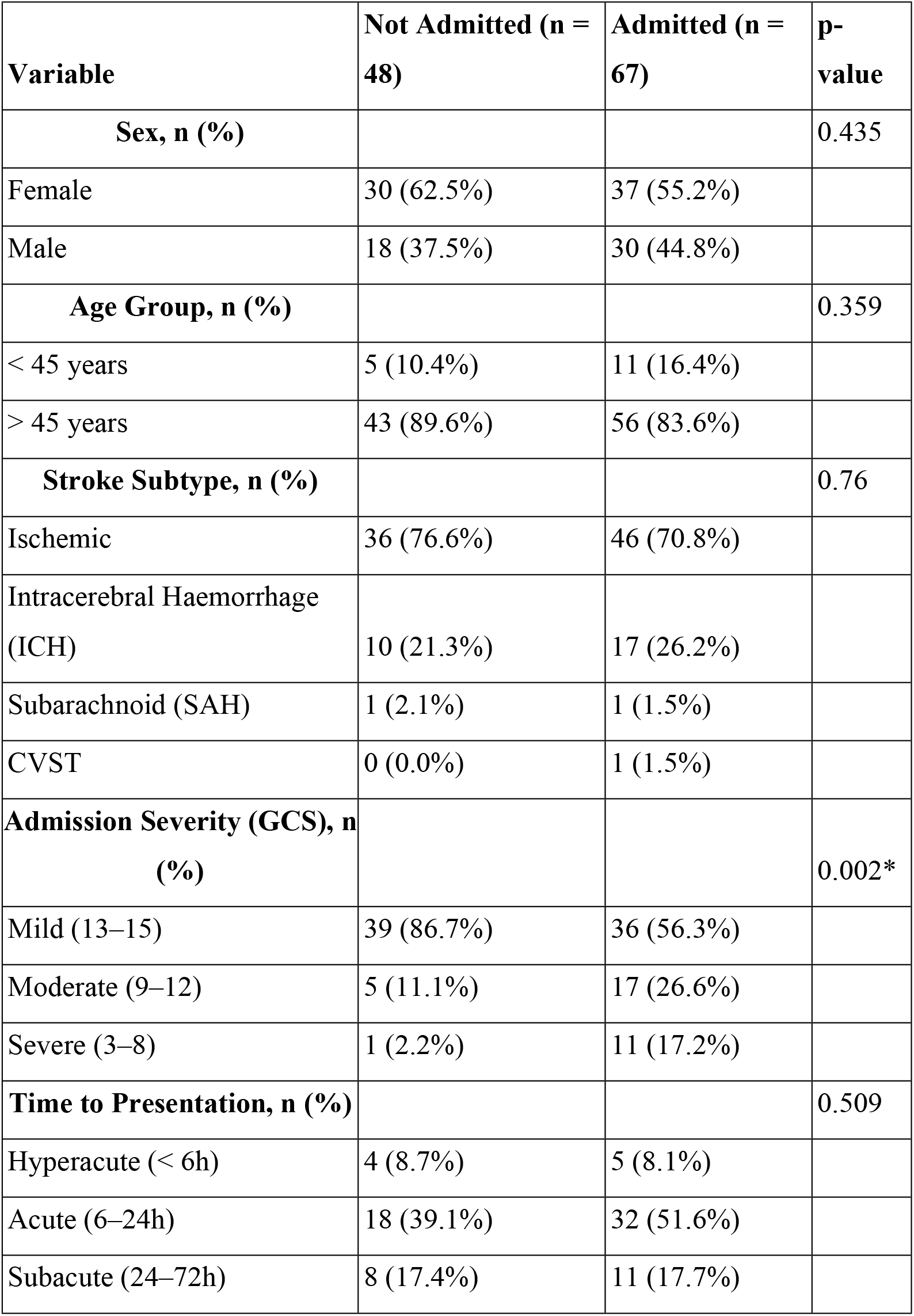
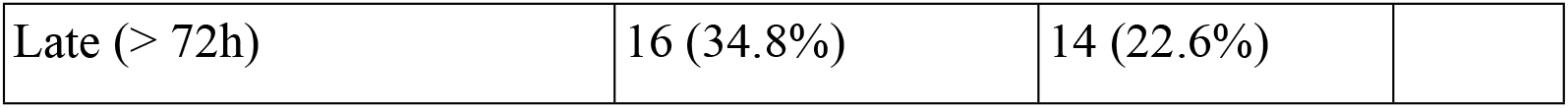
Sensitivity analysis of patients’ admission status.

## Discussion

With the limited data on stroke patterns, clinical presentations, and outcomes in Kenya, this study aimed to elicit such information while examining access to care and early rehabilitation outcomes. The findings herein provide contextually relevant data that enhances comprehension of stroke burden in Kenya and the broader SSA region.

### Stroke subtypes and demographics

The predominance of ischaemic stroke (73%) over haemorrhagic stroke (27%) in this study population mirrors global and regional from earlier estimates, which placed haemorrhagic strokes at nearly one-third of all strokes (17,22). In this study, the haemorrhagic stroke burden was still nearly double that of high-income countries, where they typically range between 10–15% (22). This disparity highlights the ongoing challenges with blood pressure control in SSA and the urgent need for context-specific prevention strategies.

### Risk factors and clinical presentation

Hypertension was the most prevalent comorbidity, aligning with global epidemiology, identifying uncontrolled blood pressure as the leading modifiable risk factor for both ischaemic and haemorrhagic strokes (23,24). Diabetic patients also constituted a significant portion (31.7%) of the cohort, with poor glycaemic control, as reflected in a mean HbA1c of 8.5%. These findings reinforce the need for integrated non-communicable disease prevention strategies tailored to the Kenyan and broader SSA context (25).

### Adherence to AHA/ASA Stroke performance measures

The audit revealed that stroke patients are managed in the general wards despite the standard recommendation to manage stroke patients in a stroke unit. Stroke units reduce mortality and disability by providing multidisciplinary and specialized care, emergency treatments, and prevention of complications (26). Regarding the standardized severity measure, the National Institute of Health Stroke Scale (NIHSS), which is the recommended severity scoring tool, was not used. This indicates a gap in measuring initial stroke severity and treatment response.

Remarkably, patients arriving within the therapeutic window for IV alteplase and thrombectomy did not receive the treatments. The facility demonstrated moderate adherence to basic secondary prevention, reflecting some awareness of the standard secondary prevention measures.

### Strengths, limitations, and future directions

A notable strength of this study lies in its implementation at a rural centre with electronic medical record infrastructure, which enabled robust and retrospective data capture. It contributes to the limited body of stroke literature from Eastern Africa, helping to fill a critical gap in regional data.

Nevertheless, the retrospective design imposes inherent limitations, including reliance on documentation accuracy and potential for missing data. Furthermore, this study did not explore longer-term functional outcomes, cognitive sequelae, or quality of life measures, which are crucial for understanding the full burden of stroke. These should be areas of focus in future prospective studies.

### Implications for practice and policy

The stark contrast between our findings and global stroke care benchmarks reinforces the need for system-wide interventions across the stroke care continuum in Kenya. From community education and early recognition to emergency referral pathways and decentralised stroke services, a multi-pronged approach is urgently needed. Particular attention should be paid to enhancing primary care-based non-communicable diseases screening and management.

Moreover, our data revealed a young and increasingly vulnerable population facing long-term stroke disability. Public health strategies should thus include targeted efforts to address the modifiable risk factors in this demographic and develop stroke prevention strategies tailored to younger populations.

Finally, investment in cardiac workup for stroke combined with the development of stroke units and specialist training, will be crucial steps toward narrowing the outcome gap between SSA and high-income countries and achieving better, more comparable stroke care outcomes.

## Data Availability

To uphold privacy and research ethics, the dataset for this study is not publicly available. However, the data may be requested through contacting the authors at

## List of abbreviations

AIC: Africa Inland Church
GCS: Glasgow Coma Scale
HIC: High Income Countries
SSA: Sub-Saharan Africa

## Ethics approval and consent to participate

Ethical approval for the study was obtained from the xxxxxxx Hospital’s Ethics Review Committee (KH/ISERC/02718/0076/2023). The patient’s data was anonymised using numbers only.

## Competing interests

None

## Funding

This work did not receive any funding.

## Author contributions

EY and PKW conceptualized the study and obtained data. EY, RWN, SK, SMG, and PKW analysed, wrote, and reviewed the manuscript.

## Acknowledgement

We are grateful to the xxxxxx Hospital management for permitting this study.

## Notes

### Competing Interest Statement

The authors have declared no competing interest.

### Author Declarations

Ethical approval for the study was obtained from the AIC Kijabe Hospital’s Ethics Review Committee (KH/ISERC/02718/0076/2023). The patient’s data was anonymised using numbers only.

